# Effectiveness of the BNT162b2 vaccine in preventing COVID-19 in the working age population – first results from a cohort study in Southern Sweden

**DOI:** 10.1101/2021.04.20.21254636

**Authors:** Jonas Björk, Malin Inghammar, Mahnaz Moghaddassi, Magnus Rasmussen, Ulf Malmqvist, Fredrik Kahn

## Abstract

**Background:** Vaccine effectiveness against COVID-19 needs to be assessed in diverse real-world population settings.

**Methods:** A cohort study of 805 741 residents in Skåne county, Southern Sweden, aged 18-64 years, of whom 26 587 received at least one dose of the BNT162b2 vaccine. Incidence rates of COVID-19 were estimated in sex- and age-adjusted analysis and stratified in two-week periods with substantial community spread of the disease.

**Results:** The estimated vaccine effectiveness in preventing infection ≥7 days after second dose was 86% (95% CI 72-94%) but only 42% (95% CI 14-63%) ≥14 days after a single dose. No difference in vaccine effectiveness was observed between females and males. Having a prior positive test was associated with 91% (95% CI 85 to 94%) effectiveness against new infection among the unvaccinated.

**Conclusion:** A satisfactory effectiveness of BNT162b2 after the second dose was suggested, but with possibly substantially lower effect before the second dose.

## Introduction

There has been a very rapid development of vaccines against SARS-CoV-2 and mass vaccination campaigns have been launched worldwide [1, 2]. To date, four different vaccines have been licensed in the European Union; BNT162b2 mRNA COVID-19 Vaccine (Pfizer-BioNTech), mRNA-1273 (Moderna Vaccine), ChAdOx1 nCoV-19 adenoviral (AstraZeneca) and Ad26.COV2-S (Jansen). In Skåne, a county in Southern Sweden with approximately 1.4 million inhabitants, the vaccination campaign started on 27 December 2020. The first to be vaccinated were nursing home residents and their caregivers as well as frontline health care workers. The aim of this study was to evaluate vaccine effectiveness (VE) of the BNT16b2 mRNA (Pfizer-BioNTech) COVID-19 vaccine in preventing SARS-CoV-2 infection in people of working age.

## Materials and methods

### Data sources

This cohort study was based on registers kept for administrative purposes at the Skåne county council, Sweden. Data sources were the total population register used for individual-level data on residency and vital status, and health care registers used for individual-level data on vaccinations and positive COVID-19 test results. Linkage between the different data sources was facilitated using the personal identification number assigned to all Swedish citizens at birth or immigration.

### Study cohort

The study cohort included all persons aged 18 – 64 years residing in Skåne county, Sweden, on 27 December 2020 when vaccinations started. The cohort was followed until 28 February 2021. Data on vaccination, type of vaccine and dose, were linked to the cohort, together with data on prior positive COVID-19 tests at any time point from March 2020 until 26 December 2020. Individuals who during follow up were vaccinated with other COVID-19 vaccines than BNT16b2 mRNA were excluded at baseline due to too small numbers to permit evaluation (1.0 % of the population). Individuals moving out from the region during follow up were censored on the date of relocation.

### Outcomes

The primary outcome was the first positive SARS-CoV2 test result received from December 27 2020 to February 28 2021, hereafter called COVID-19-infection. During the study period, the Regional Center for Disease Control recommended individuals of >6 years old with symptoms of COVID-19 to get tested. Additionally, test recommendations were from January 21 2021 given to persons living in the same household as a person with a confirmed infection, irrespective of own symptoms, five days after the index case. Sampling was performed mainly from nasopharynx and analysed by RT-PCR at the Regional Laboratory of Clinical Microbiology or through a combined sampling from pharynx, nose and saliva through RT-PCR at laboratories assigned by the Swedish Board of Health and Welfare: Dynamic Code AB, Linköping, Sweden and Eurofins LifeCodexx GmbH, Germany. Moreover, some patients and health care workers were tested using antigen tests (PANBIO™, Abbot) from nasopharynx samples, within both primary and secondary care. Result from all diagnostic modalities and laboratories were available for the study. As secondary outcome, we used death in COVID-19, defined as death within 30 days of a positive test.

### Statistical analysis

Statistical analyses were conducted in Stata SE 14.2 (Stata Corp.) and IBM SPSS Statistics 26 (SPSS Corp.). The number of the positive COVID-19 tests was calculated in relation to person-weeks of follow up, separately for unvaccinated and vaccinated follow up time, and stratified on prior COVID-19 positivity. Further stratifications were done according to i) no dose or 0 - 13 days after the first dose, ii) at least 14 days after the first dose but before second dose, iii) 0 - 6 days after the date of the second dose, iv) at least 7 days after the second dose. To account for variations in community spread during follow up, the counting of cases and person-weeks was done separately in four two-week periods (period 1: Dec 27 –Jan 17, period 2: Jan 18 – 31, period 3: Feb 1 – 14 and period 4: Feb 15 – 28). We estimated the VE overall and stratified by sex among individuals with no prior positive test at baseline as (*IRR* – 1) / IRR together with 95% confidence interval (CI), where IRR represents the incidence rate ratio contrasting unvaccinated with vaccinated person-time. Main VE results were reported for period 4 with the longest follow up, but we also present results for period 1-3 as comparison. As a further reference, we calculated effectiveness associated with a prior positive test at baseline. All statistical analyses were weighted to account for differences in sex and age distribution (five groups: 18 – 44, 45 – 49, 50 – 54, 55 – 59 and 60 – 64 years old) among vaccinated and unvaccinated.

## Results

The study cohort comprised 805 741 individuals on 27 December 2020, of whom 26 587 (3.3%) received at least one dose of the BNT16b2 mRNA vaccine until 28 February 2021 (Table 1). The vaccinated cohort had a higher proportion of females (80% vs. 52%) and was older (median age 47 vs. 40 years) than the unvaccinated cohort. The estimated VE in preventing infection 7 days or more after second dose among subjects with no prior positive test was 86% (95% CI 72 to 94%) during period 4 (Feb 15-28; Table 2 and Figure 1). Similar but more statistically uncertain VE (93%; 95% CI 59 to 100%) was observed in period 3 (Figure 1), whereas the VE after second dose could not be evaluated in period 1-2 (Table S1). VE was similar among females and males, but more statistically uncertain among males due to fewer vaccinated (Table S2). No deaths within 30 days of a positive test were observed among the vaccinated (Table S3) Having a prior positive test was associated with 91% (95% CI 85 to 94%) effectiveness against new infection among the unvaccinated during period 4 (Table 2). This protective effect was similarly high during period 3 (Table S1), and still high when restricting the analysis to individuals with a prior positive test more than three months before baseline (83%, 95% CI 51 to 97%; not in tables).

**Table 1.** Baseline characteristics of the study cohort on 27 December 2020 when COVID-19 vaccination started, stratified by vaccination status 28 February 2021.

|  | Vaccinated<br>(n = 26 587) | Unvaccinated<br>(n = 779 154) | Uptake (% of total)<br>(3.3) |
| --- | --- | --- | --- |
| Sex, n (%) |  |  |  |
| Female | 21 274 (80.0) | 404 808 (52.0) | 5.4 |
| Male | 5 313 (20.0) | 374 346 (48.0) | 1.3 |
| Age by sex, n (%) |  |  |  |
| Female, 18 – 44 | 8 947 (33.7) | 223 877 (28.7) | 3.8 |
| 45 – 49 | 2 711 (10.2) | 40 405 (5.2) | 6.3 |
| 50 – 54 | 3 097 (11.6) | 39 870 (5.1) | 7.2 |
| 55 – 59 | 3 572 (13.4) | 37 283 (4.8) | 8.7 |
| 60 – 64 | 2 947 (11.1) | 32 911 (4.2) | 8.2 |
| Male, 18 – 44 | 2 854 (10.7) | 239 332 (30.7) | 1.2 |
| 45 – 49 | 541 (2.0) | 44 580 (5.7) | 1.2 |
| 50 – 54 | 554 (2.1) | 43 876 (5.6) | 1.2 |
| 55 – 59 | 644 (2.4) | 41 427 (5.3) | 1.5 |
| 60 – 64 | 720 (2.7) | 35 593 (4.6) | 2.0 |
| Country of birth |  |  |  |
| Sweden | 20 725 (78.0) | 547 873 (70.3) | 3.6 |
| Abroad | 5 862 (22.0) | 231 281 (29.7) | 2.5 |
| Civil status |  |  |  |
| Unmarried | 10 088 (37.9) | 380 434 (48.8) | 2.6 |
| Married | 12 066 (45.4) | 307 642 (39.5) | 3.8 |
| Divorced | 4 065 (15.3) | 85 723 (11.0) | 4.5 |
| Widow/widower | 368 (1.4) | 5 355 (0.7) | 6.4 |
| Positive SARS-CoV-2 test before baseline | 2 660 (10.0) | 36 740 (4.7) | 6.8 |

**Table 2.** Effectiveness of the BNT16b2 mRNA (Pfizer-BioNTech) vaccine in preventing SARS-CoV-2 infection during period 4 (15 – 28 February 2021).

|  | Cases, n | Person-time, weeks | Incidence (95% CI)<sup>a</sup> | Effectiveness, % (95% CI) |
| --- | --- | --- | --- | --- |
| <u>No prior positive test</u> |  |  |  |  |
| Unvaccinated or before 1<sup>st</sup> dose | 4 155 | 1 414 660 | 294 (285 – 303) | Ref. |
| 1<sup>st</sup> dose, day 0-13 | 9 | 2 260 | 398 (138 – 658) | Not evaluated |
| 1<sup>st</sup> dose, day 14- | 25 | 14 690 | 170 (103 – 237) | 42 (14 – 63) |
| 2<sup>nd</sup> dose, day 0-6 | 10 | 8 537 | 117 (45 – 190) | 60 (27 – 81) |
| 2<sup>nd</sup> dose, day 7- | 8 | 19 088 | 42 (13 – 71) | 86 (72 – 94) |
| <u>Prior positive test<sup>b</sup></u> |  |  |  |  |
| Unvaccinated or before 1<sup>st</sup> dose | 21 | 75 399 | 28 (16 – 40) | 91 (85 – 94) |

<sup>a</sup>Cases per 100 000 persons and week (95% confidence interval). Results from statistical analysis were weighted with respect to sex and age distribution of the vaccinated cohort.
<sup>b</sup>The number of vaccinated with prior positive test was too few to permit evaluation of vaccine effectiveness

**Figure 1.**
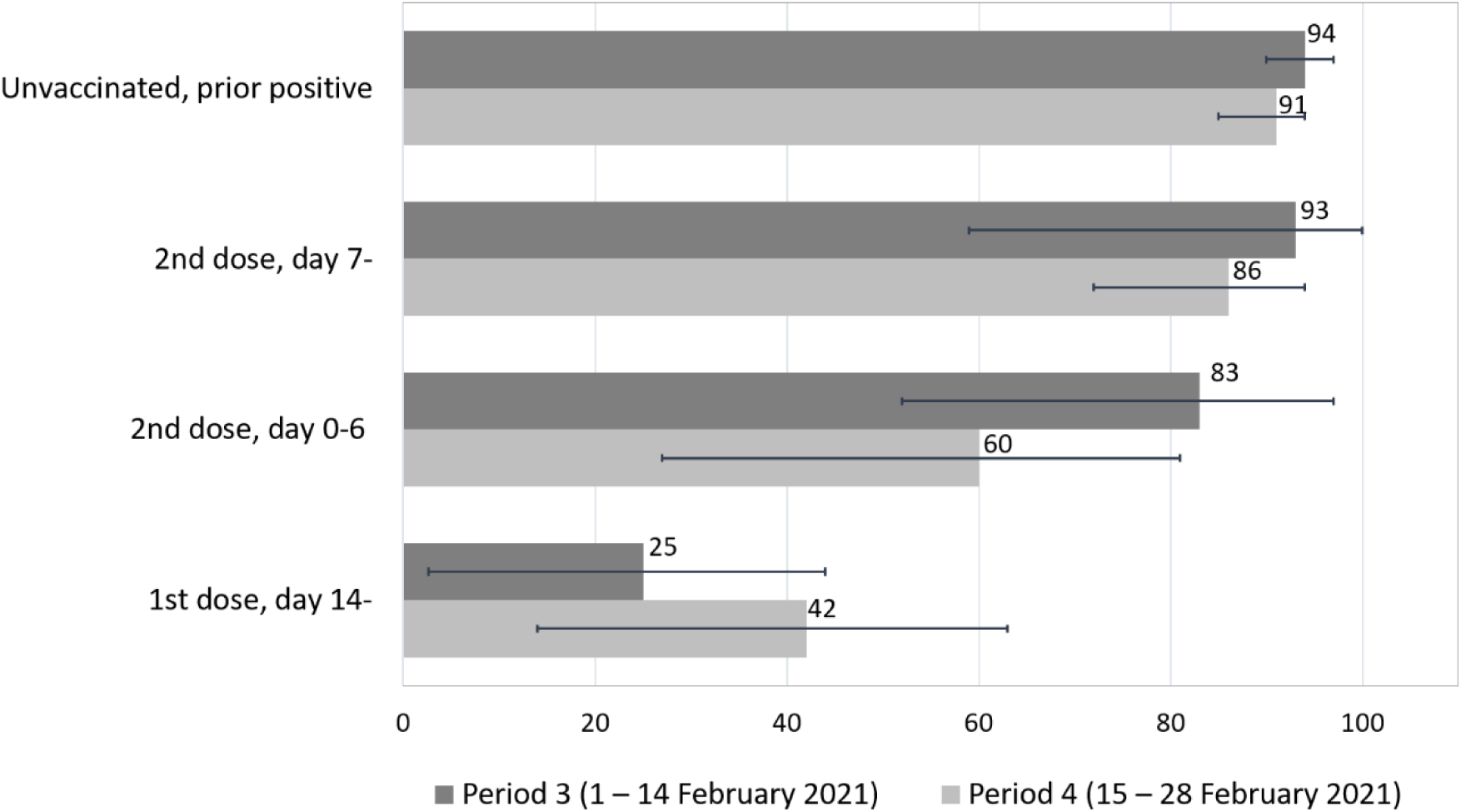
Effectiveness of the BNT16b2 mRNA (Pfizer-BioNTech) vaccine in preventing SARS-CoV-2 infection during period 3 (1 – 14 February 2021) and 4 (15 – 28 February 2021).

## Discussion

The most salient finding was the satisfactory VE in preventing SARS-CoV-2 infection seven days or more after the second dose of the BNT16b2 mRNA vaccine, observed in a working age population. A major strength of the study was the rapid evaluation of vaccine effectiveness in a real-world Scandinavian setting with substantial and prevailing community spread of the virus. The circulation of SARS-CoV-2 in the region was among the highest in Europe during the study period with incidence rates between 300 and 900 new cases per week and 100 000 population, why the vaccinated cohort most likely had considerable exposure to the virus during follow up.

A limitation was the short follow up time, and the current lack of data to evaluate effects on disease severity and hospitalizations and effects of specific virus variants. Surveillance data compiled by the Public Health Agency of Sweden suggest that 32 – 50% of the positive tests were of the B.1.1.7 variant in the study region during the last follow up period [3]. We also lacked data on disease history and co-existing conditions in the study population, preventing a detailed matching of vaccinated and unvaccinated beyond sex, age and follow up period. The main reason for vaccination in the study cohort was working in the health care sector, but individuals aged up to 64 years who were vaccinated due to their residence in special homes were also included. As we could not account for differences in health related to occupational status and residence across cohorts, we decided not to evaluate effects on all-cause mortality. However, we observed no deaths related to COVID-19 among the vaccinated. As a final limitation, it should be noted that we may to some extent underestimate VE due to unknown prior infections, especially as COVID-19 testing was limited in this population during the spring 2020.

Several reports on VE of the BNT162b2 mRNA vaccine have already emerged since the recent launch of large vaccination campaigns in many parts of the world, Although we estimated the VE after 14 days after the first dose, we also studied the effect 0-6 days after the second dose with a comparably low estimated VE (60%) where the effect is probably still due to the first dose. A cohort study in health care workers in UK demonstrated a VE against COVID-19 infection after first dose that was higher than in our study (72% after 21 days), whereas they found similar VE as we after second dose (86% after 7 days) [4]. Other studies have also reported higher VE after the first dose [5, 6], and reduced risk of severe COVID-19 that required hospitalization [7]. However, a cohort study from Israel with detailed matching on demographic and clinical characteristics in a diverse population showed similar evolvement of VE after first and second dose as in our study when evaluated against symptomatic infection (57% 14 - 20 days after first dose and 94% 7 days after second dose [8].

The suggested high protection (91%-94% depending on level of community spread) by a previous infection in our study is in line with recently published studies. A study from Denmark suggested an overall protection against reinfection of 81% during the second surge of the COVID-19 epidemic, but with markedly diminishing protection of individuals ≥65 years old [9]. Among health care workers in UK the estimated protection of a previous infection was 94% against a probable or possible symptomatic infection and 83% against all probable and possible infections (our calculations based on reported odds ratios) [10].

As our results suggest that vaccine effectiveness may not be satisfactory until seven days after the second dose, it is prudent to inform the public about the importance of maintaining social distancing and complying with other recommendations until full vaccine effect can be expected. Compliance with recommendations is likely to be especially important in regions where the exposure to the virus is still considerable. Another aspect of the present findings, especially when making priorities in the vaccination programs for the general population, is the strong protective effect associated with documented prior infection. It is important to continue to monitor VE for longer periods and to compare VE of different vaccines, and also carefully monitoring risk of adverse events. Sweden, with its combination of register infrastructure for population studies and prevailing community spread of the SARS-CoV-2 virus, constitutes a suitable setting for such further studies.

## Conclusion

In conclusion, we found a vaccine effectiveness of 86% in preventing infection 7 days or more after second dose of BNT16b2 mRNA vaccine, in adults of working age during a period of high circulation of SARS-CoV-2. The observed vaccine effectiveness was not satisfactory after a first dose only.

## Data Availability

The dataset used in the present study is hosted by Scania county council, Sweden. Legal and ethical
restrictions prevent public sharing of the dataset. Data can be made available for collaborations upon request to interested researchers but would generally require a new ethical permission.

## Acknowledgments

Cecilia Åkesson-Kotsaris, Paul Söderholm and Helena Hallefjord, Clinical Studies Sweden, for excellence in bringing the data infrastructure in place. Susann Ullén, Clinical Studies Sweden, for statistical advice.

## Author contribution

All authors conceived and designed the study, analyzed and interpreted the results. UM, JB and MM acquired data, JB and MM conducted the statistical analyses. JB, FK, and MI drafted the manuscript. All authors critically revised the manuscript and approved the final version for submission. FK supervised the study and is the guarantor.

## Conflicts of interest

All authors declare no conflicts of interest, no support or financial relationship with any organization or other activities with any influence on the submitted work.

## Funding

This study was supported by an internal grant for thematic collaboration initiatives at Lund University held by JB, and by Swedish Research Council (VR; grant number 2019-00198). FK is supported by grants from the Swedish Research Council and Governmental Funds for Clinical Research (ALF). The funders played no role in the design of the study, data collection or analysis, decision to publish, or preparation of the manuscript.

## Ethics and Permissions

Ethical approval was obtained from the Swedish Ethical Review Authority (2021-00059).

## Supporting information

**Table S1**. Effectiveness of the BNT16b2 mRNA (Pfizer-BioNTech) vaccine in preventing SARS-CoV-2 infection during period 1-3 (27 December 2020 – 14 February 2021).

**Table S2**. Effectiveness of the BNT16b2 mRNA (Pfizer-BioNTech) vaccine in preventing SARS-CoV-2 infection during period 4 (15 – 28 February 2021) and stratified by sex.

**Table S3**. Effectiveness of the BNT16b2 mRNA (Pfizer-BioNTech) vaccine on COVID-19 mortality during follow up 27 December 2020 – 28 February 2021.

## Notes

### Competing Interest Statement

The authors have declared no competing interest.

### Author Declarations

Ethical approval was obtained from the Swedish Ethical Review Authority (2021-00059). As the study is register-based, individual participant consent was not necessary.

