## Supporting information for "Effectiveness of the BNT162b2 vaccine in preventing COVID-19 in the working age population – first results from a cohort study in Southern Sweden"

**Table S1.** Effectiveness of the BNT16b2 mRNA (Pfizer-BioNTech) vaccine in preventing SARS-CoV-2 infection during period 1-3 (27 December 2020 – 14 February 2021).

**Table S2.** Effectiveness of the BNT16b2 mRNA (Pfizer-BioNTech) vaccine in preventing SARS-CoV-2 infection during period 4 (15 – 28 February 2021) and stratified by sex.

**Table S3.** Effectiveness of the BNT16b2 mRNA (Pfizer-BioNTech) vaccine on COVID-19 mortality during follow up 27 December 2020 – 28 February 2021.

**Table S1.** Effectiveness of the BNT16b2 mRNA (Pfizer-BioNTech) vaccine in preventing SARS-CoV-2 infection during period 1-3 (27 December 2020 – 14 February 2021).

| | 27 Dec 2020 – 17 Jan 2021 | | | 18 – 31 Jan 2021 | | | 1 – 14 Feb 2021 | | | |
|---|---|---|---|---|---|---|---|---|---|---|
| | Cases, n | Person-time, weeks | Incidence (95% CI)[a] | Cases, n | Person-time, weeks | Incidence (95% CI)[a] | Cases, n | Person-time, weeks | Incidence (95% CI)[a] | Effectiveness, % (95% CI) |
| No prior positive test | | | | | | | | | | |
| Unvaccinated or before 1st dose | 20 215 | 2 257 089 | 896 (883 – 908) | 6 885 | 1 451 061 | 474 (463 – 486) | 4 787 | 1 426 823 | 336 (326 – 345) | Ref. |
| 1st dose, day 0-13 | 51 | 7 088 | 720 (522 – 917) | 176 | 21 884 | 804 (685 – 923) | 46 | 10 795 | 426 (303 – 549) | - |
| 1st dose, day 14- | 3 | 488 | 615 (127 – 1 797) | 22 | 7 711 | 285 (166 – 405) | 55 | 21 952 | 251 (184 – 317) | 25 (2.6 – 44) |

| | | | | | | | | | | |
|---|---|---|---|---|---|---|---|---|---|---|
| 2nd dose, day 0-6 | - | - | - | - | - | - | 3 | 5 397 | 56 (11 – 162) | 83 (52 – 97) |
| 2nd dose, day 7- | - | - | - | - | - | - | 1 | 4 078 | 25 (0.6 – 137) | 93 (59 – 100) |
| <u>Prior positive test</u> [b] | | | | | | | | | | |
| Unvaccinated or before 1st dose | NR | NR | NR | 21 | 77 288 | 28 (16 – 39) | 15 | 75 767 | 19 (9 – 29) | 94 (90 – 97) |

[a]Cases per 100 000 persons and week (95% confidence interval). Results from statistical analysis were weighted with respect to sex and age distribution of the vaccinated cohort.

[b]The number of vaccinated with prior positive test was too few to permit evaluation of vaccine effectiveness

**Table S2.** Effectiveness of the BNT16b2 mRNA (Pfizer-BioNTech) vaccine in preventing SARS-CoV-2 infection during period 4 (15 – 28 February 2021) and stratified by sex.

| | Females | | | | Males | | | |
|---|---|---|---|---|---|---|---|---|
| | Cases, n | Person-time, weeks | Incidence (95% CI)[a] | Effectiveness, % (95 CI) | Cases, n | Person-time, weeks | Incidence (95% CI)[a] | Effectiveness, % (95 CI) |
| No prior positive test | | | | | | | | |
| Unvaccinated or before 1st dose | 3 313 | 1 129 981 | 293 (283 – 303) | Ref. | 842 | 285 208 | 295 (275 – 315) | Ref. |
| 1st dose, day 0-13 | 8 | 15 914 | 503 (154 – 851) | - | 1 | 669 | 149 (3.8 – 833) | - |
| 1st dose, day 14- | 22 | 11 738 | 187 (109 – 266) | 36 (3.0 – 60) | 3 | 2 952 | 102 (21 – 297) | 66 (0.0 – 93) |

| | | | | | | | | |
|---|---|---|---|---|---|---|---|---|
| 2nd dose, day 0-6 | 9 | 7 016 | 128 (44 – 212) | 56 (17 – 80) | 1 | 1 521 | 66 (17 – 366) | 78 (0.0 – 99) |
| 2nd dose, day 7- | 6 | 15 397 | 39 (8 – 70) | 87 (71 – 95) | 2 | 3 690 | 54 (6.6 – 196) | 82 (33 – 98) |
| Prior positive test[b] | | | | | | | | |
| Unvaccinated or before 1st dose | 20 | 62 265 | 31 (18 – 45) | 89 (83 – 93) | 1 | 13 134 | 7.6 (0.2 – 42) | 97 (86 – 100) |

[a]Cases per 100 000 persons and week (95% confidence interval). Results from statistical analysis were weighted with respect to sex and age distribution of the vaccinated cohort.

[b]The number of vaccinated with prior positive test was too few to permit evaluation of vaccine effectiveness

**Table S3.** Effectiveness of the BNT16b2 mRNA (Pfizer-BioNTech) vaccine on COVID-19 mortality during follow up 27 December 2020 – 28 February 2021.

| | Deaths | Person-time, weeks | Mortality rate (95% CI)[a] |
|---|---|---|---|
| Unvaccinated or before 1st dose | 36 | 7 102 506 | 0.5 (0.3 – 0.7) |
| 1st dose, day 0-13 | 0 | 48 257 | 0.0 (0.0 – 7.6) |
| 1st dose, day 14- | 0 | 52 102 | 0.0 (0.0 – 7.1) |
| 2nd dose, day 0-6 | 0 | 16 544 | 0.0 (0.0 – 22.0) |
| 2nd dose, day 7- | 0 | 26 725 | 0.0 (0.0 – 13.8) |

[a]Deaths per 100 000 persons and week (95% confidence interval). Results from statistical analysis were weighted with respect to sex and age distribution of the vaccinated cohort.
